# Geographical surveillance of COVID-19: Diagnosed cases and death in the United States

**DOI:** 10.1101/2020.05.22.20110155

**Authors:** Raid Amin, Terri Hall, Jacob Church, Daniela Schlierf, Martin Kulldorff

**Affiliations:** Department of Mathematics and Statistics, University of West Florida, Pensacola, FL 32514, USA; Division of Pharmacoepidemiology and Pharmacoeconomics, Department of Medicine, Harvard Medical School and Brigham and Women’s Hospital, Boston, MA 02120, USA

**Keywords:** Clusters, prospective space-time analysis, spatial analysis, COVID-19

## Abstract

**Background:** COVID-19 is a new coronavirus that has spread from person to person throughout the world. Geographical disease surveillance is a powerful tool to monitor the spread of epidemics and pandemic, providing important information on the location of new hot-spots, assisting public health agencies to implement targeted approaches to minimize mortality.

**Methods:** County level data from January 22-April 28 was downloaded from USAfacts.org to create heat maps with ArcMap™ for diagnosed COVID-19 cases and mortality. The data was analyzed using spatial and space-time scan statistics and the SaTScan™ software, to detect geographical cluster with high incidence and mortality, adjusting for multiple testing. Analyses were adjusted for age. While the spatial clusters represent counties with unusually high counts of COVID-19 when averaged over the time period January 22-April 20, the space-time clusters allow us to identify groups of counties in which there exists a significant change over time.

**Results:** There were several statistically significant COVID-19 clusters for both incidence and mortality. Top clusters with high rates included the areas in and around New York City, New Orleans and Chicago, but there were also several small rural clusters. Top clusters for a recent surge in incidence and mortality included large parts of the Midwest, the Mid-Atlantic Region, and several smaller areas in and around New York and New England.

**Conclusions:** Spatial and space-time surveillance of COVID-19 can be useful for public health departments in their efforts to minimize mortality from the disease. It can also be applied to smaller regions with more granular data.

## Background

The COVID-19 pandemic is closely followed around the world, with daily estimates of diagnosed case counts and death for almost every country in the world. [1][2][3] Regional and local numbers are also important. Our focus here is on the cluster analysis methodology used to provide a comprehensive and effective disease surveillance of COVID-19 with which it is possible to summarize results in a clear and meaningful manner where the hotspots (clusters) are located, and to identify where such clusters are located and whether some clusters seem to be emerging or spreading. While descriptive maps of COVID-19 cases or death counts provide useful information on the disease, it can be hard to distinguish random variation from true hotspots. Spatial and space-time scan statistics can detect groups of neighboring counties with high rates of COVID-19, and determine whether they are statistically significant. The use of heat maps complemented by scan statistics allows us to better understand the geographic distribution of COVID-19 across the contiguous USA.

There are various reasons for evaluating the geographical distribution of COVID-19, and each requires a different type of scan statistic. In this paper, we give examples of each type of question and its corresponding scan statistic.

COVID-19 will continue to be a major threat to older people until herd immunity arrives, either through natural disease or a potential future vaccine. In order to understand geographical differences in movement along the path towards herd immunity, we can use a purely spatial scan statistic and the cumulative number of diagnoses or deaths as a proportion of the population. While it would be better to use large-scale random antibody testing, that is not yet available.

With cumulative data, it is not possible to distinguish areas with a currently high disease incidence versus areas where things used to be bad but where the spread has subsided. For an evaluation of the current situation, it is instead appropriate to only analyze the last one, two or three weeks of data, while still employing the purely spatial scan statistic.

A purely spatial analysis with only a few weeks of data cannot distinguish high incident areas that have been high for a while from areas where the incidence is increasing. The detection of the latter is interesting even if they are not yet as high as many other parts of the country. Such analyses can be done using all the cumulative data with a prospective space-time scan statistic, adjusting for geographical differences. In essence, each geographical area is evaluated as to whether the incidence or mortality rate has increased compared to what it used to be in that same area.

A fourth option is to do a prospective space-time analysis without adjusting for geography, as performed by. Desjardins et al [4]. Such an analysis detects areas that are high during recent days or weeks, when compared to the national average in both current and past time, while it can but does not have to be higher than the prior rates in the same area. The purpose of such an analysis is hence different from a space-time adjusted analysis that adjusts for geographical variation, and the two types of analysis complements each other.

In order to illustrate how such a comprehensive set of methods can be applied, we used COVID-19 data for US counties, to evaluate the geographical distribution of infection cases and also for deaths associated with COVID-19. We created informative heat maps with superimposed cluster rings that identify significant clusters from SaTScan™ based on the Poisson distribution. [5]

## Materials and Methods

### COVID-19 Data

COVID-19 data were obtained from the not-for-profit organization USA facts, at www.usafacts.org. [6] They have in turn obtained the data from the Centers for Disease Control and Prevention. [7] as well as from state and county level public health agencies. Confirmed diagnosed cases and deaths are given as cumulative counts starting January 22, 2020 with a daily update. Since only confirmed diagnoses are used, many cases are missed and the proportion of missed cases will depend on the amount of testing conducted, which may vary geographically.

For the purely spatial analyses we used data for January 22-April 20 and the two week period of April 7 – April 20. For the space-time analysis we used data for January 22-April 28.

### Age adjustment

COVID-19 diagnoses and deaths vary greatly by age, and since counties have different age distribution, it is important to adjust the spatial analyses for age. For each county, we used the age-specific data from the Census Bureau.[8] For COVID-19 diagnoses and deaths, we used the age-distribution reported by CDC.[9] Age was adjusted for using indirect standardization

### Choropleth Maps

For descriptive purposes, choropleth maps were constructed. For the 3,108 counties, the age-adjusted rates were ranked from lowest to highest and sorted into quintiles. In the figures, each county is colored from dark green (lowest COVID-19 rates), to light green, yellow, orange, and dark red (highest COVID-19 rates). The legend gives the range of values for each shade of color, which is different for different maps. This type of coloring allows the reader to quickly scan the US map and to eyeball where parts of the USA have high or low rates of this virus.

### The Spatial Scan Statistic

To detect geographical areas with a statistically significant excess number of COVID-19 diagnoses or deaths, we used the spatial scan statistic, a widely used method for geographical disease surveillance. This method detects and determines statistical significance of geographical cluster without having to pre-specify the cluster size or location, while automatically adjusting for the multiple testing that exists in the large number of potential clusters evaluated.

The spatial scan statistic applies a moving circular window on the map, centered on each of many possible grid points positioned throughout the study region. County centroids were used for the grid points to ensure that each county could be a potential cluster by itself. For each grid point, the radius of the window is varied continuously in size from zero up to some upper limit that we set to 10% of the total population at risk. This way the circular window is flexible both in location and size. Overall, this method creates many thousands of distinct geographical circles with different sets of neighboring counties within them. Each circle is a possible candidate for a cluster.

Using the observed and age-adjusted expected counts for each county, the Poisson distribution was used to model the random nature of the counts. P-values were calculated by fitting the Gumbel distribution to 999 random data sets generated under the null hypothesis of equal risk throughout the country. [10]

### The Prospective Space-Time Scan Statistic

To detect recent increases, a prospective space-time cluster analysis was performed for both COVID-19 cases and for mortality. A nonparametric spatial adjustment was used in order to detect areas with localized temporal increases, while not comparing rates across geographical regions. This means that a cluster can be detected if there is a temporal increase in the area even if that area still has a lower rate than the rest of the country.

The prospective space-time scan statistic identifies newly emerged clusters by using a cylindrical three-dimensional scanning window. The circular base is the same as for the purely spatial scan statistic while the height of the cylinder corresponds to time. For each circular location and size, different time lengths are evaluated up to a maximum of 14 days. In the prospective version used here, only cylinders that include the last day of the study period are used, as we are only interested in emerging clusters but not historical ones. The prospective space-time analysis the scan statistic can be used for time-periodic surveillance in which the analysis is repeated each week. [10][11]

All analyses were conducted using the free SaTScan™ software (www.satscan.org). [5]

## Results

### Cumulative Data

For January 22-April 20, 2020, the geographical distribution of confirmed COVID-19 diagnoses are depicted in Figure 1, with 38 statistically significant clusters (p<0.05). Table 1 provides detailed information on the top 20 clusters with the smallest p-values. The top cluster was in and around New York City, with over ten times as many diagnosed cases compared to the rest of the country. Cluster 2 is in and around New Orleans, Louisiana, with relative risk (RR) 5.7. More unexpectedly, there are several small clusters with very high relative risk, including Marion County, Ohio; Blaine County, Idaho; Lincoln County, Arkansas; and Louisa County, Iowa.

**Figure 1:**
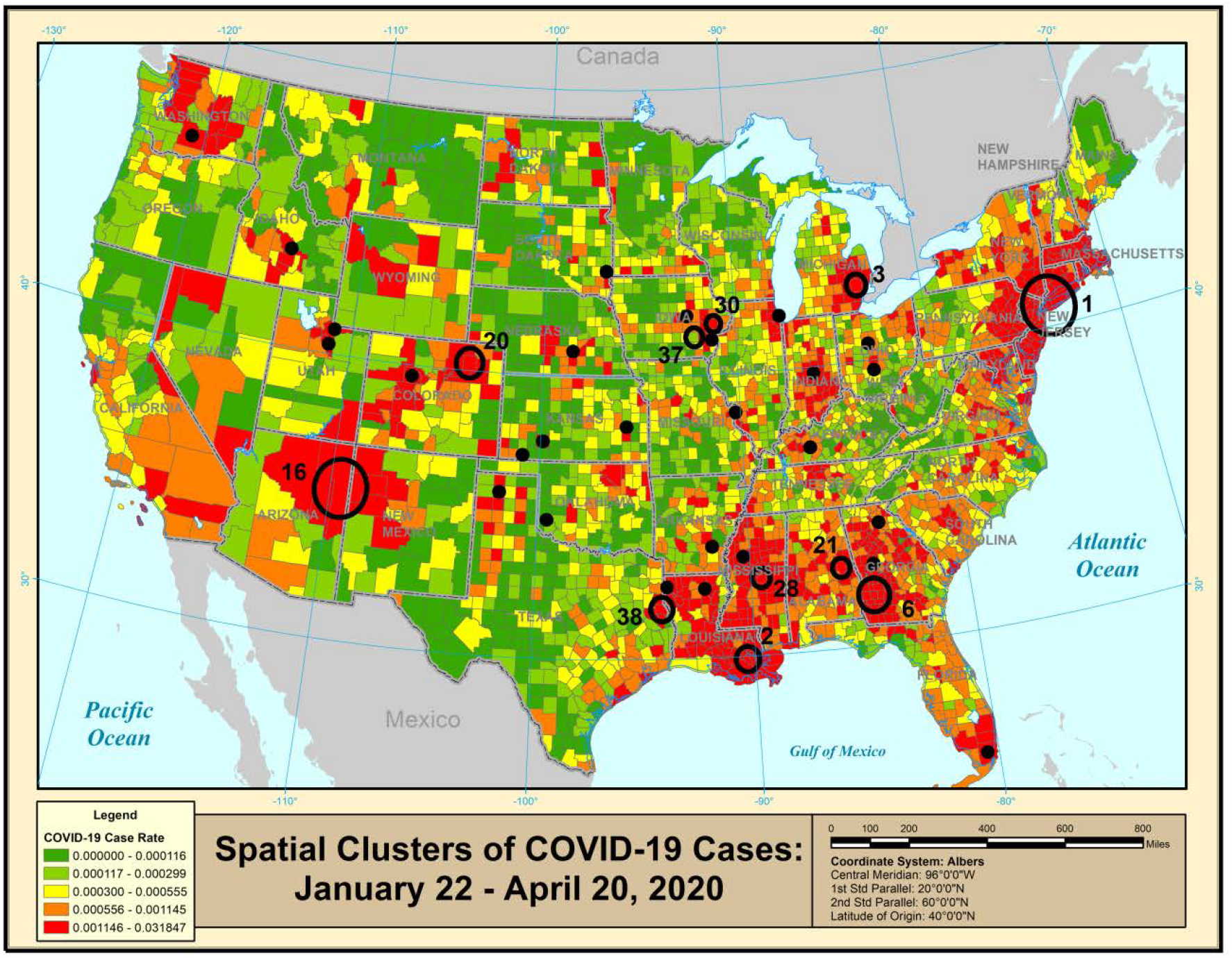
Geographical distribution and clusters of diagnosed COVID-19 cases for January 22-April 20, 2020.

**Table 1:**
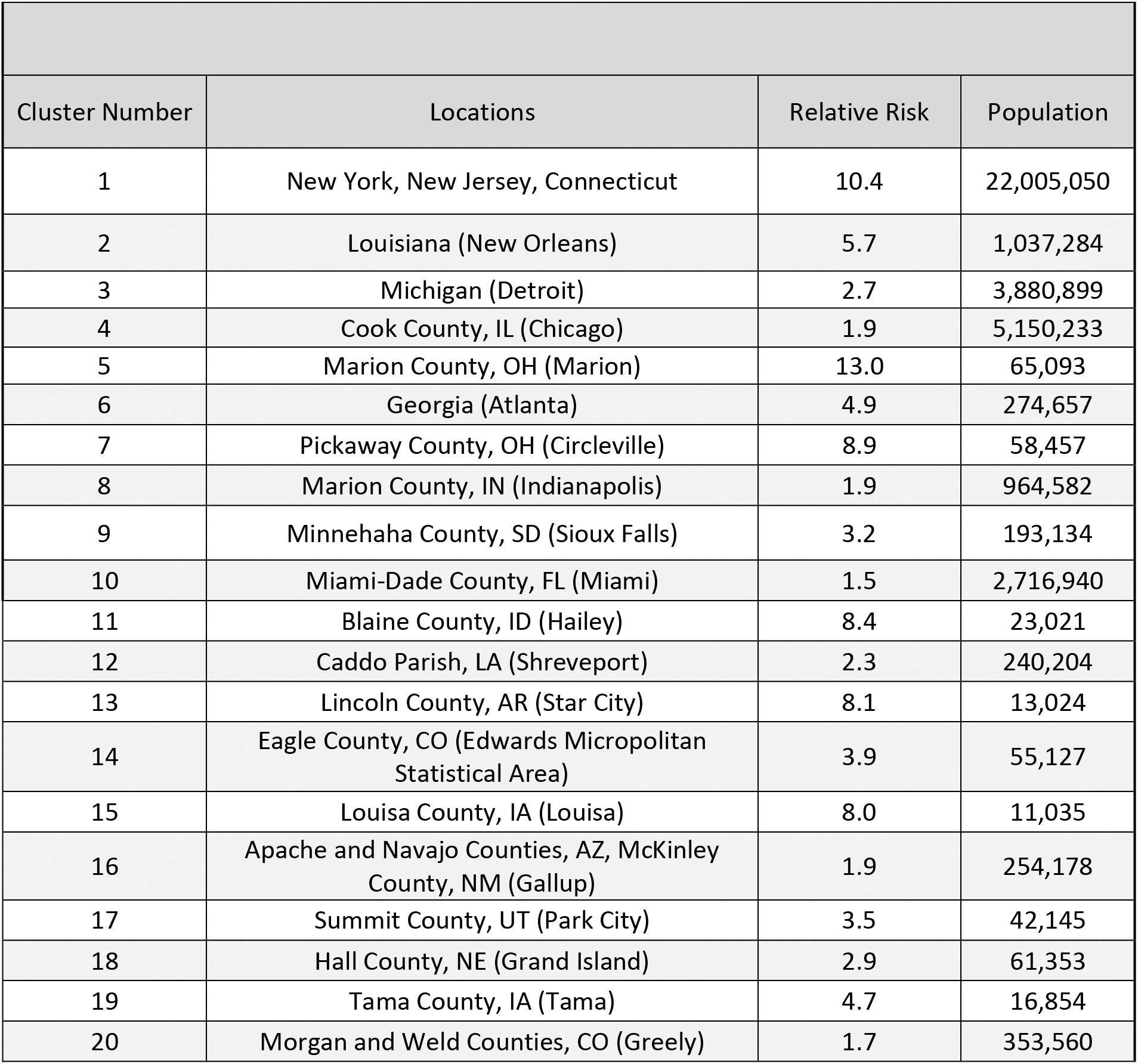
Statistically significant spatial clusters (p<0.05) of diagnosed COVID-19 cases for January 22-April 20, 2020, adjusted for age and multiple testing.

| Cluster Number | Locations | Relative Risk | Population |
| --- | --- | --- | --- |
| 1 | New York, New Jersey, Connecticut | 10.4 | 22,005,050 |
| 2 | Louisiana (New Orleans) | 5.7 | 1,037,284 |
| 3 | Michigan (Detroit) | 2.7 | 3,880,899 |
| 4 | Cook County, IL (Chicago) | 1.9 | 5,150,233 |
| 5 | Marion County, OH (Marion) | 13.0 | 65,093 |
| 6 | Georgia (Atlanta) | 4.9 | 274,657 |
| 7 | Pickaway County, OH (Circleville) | 8.9 | 58,457 |
| 8 | Marion County, IN (Indianapolis) | 1.9 | 964,582 |
| 9 | Minnehaha County, SD (Sioux Falls) | 3.2 | 193,134 |
| 10 | Miami-Dade County, FL (Miami) | 1.5 | 2,716,940 |
| 11 | Blaine County, ID (Hailey) | 8.4 | 23,021 |
| 12 | Caddo Parish, LA (Shreveport) | 2.3 | 240,204 |
| 13 | Lincoln County, AR (Star City) | 8.1 | 13,024 |
| 14 | Eagle County, CO (Edwards Micropolitan Statistical Area) | 3.9 | 55,127 |
| 15 | Louisa County, IA (Louisa) | 8.0 | 11,035 |
| 16 | Apache and Navajo Counties, AZ, McKinley County, NM (Gallup) | 1.9 | 254,178 |
| 17 | Summit County, UT (Park City) | 3.5 | 42,145 |
| 18 | Hall County, NE (Grand Island) | 2.9 | 61,353 |
| 19 | Tama County, IA (Tama) | 4.7 | 16,854 |
| 20 | Morgan and Weld Counties, CO (Greely) | 1.7 | 353,560 |

For COVID-19 deaths, there are ten significant clusters for the same time period, as shown in Figure 2 and Table 2. Again, the top cluster is in the New York-New Jersey-Connecticut area, with nearly 21 million people, and with a COVID-19 mortality that is 14 times higher than the rest of the United States. In general, results are similar for diagnoses and deaths, although the latter analysis has fewer clusters due to a smaller sample size.

**Figure 2:**
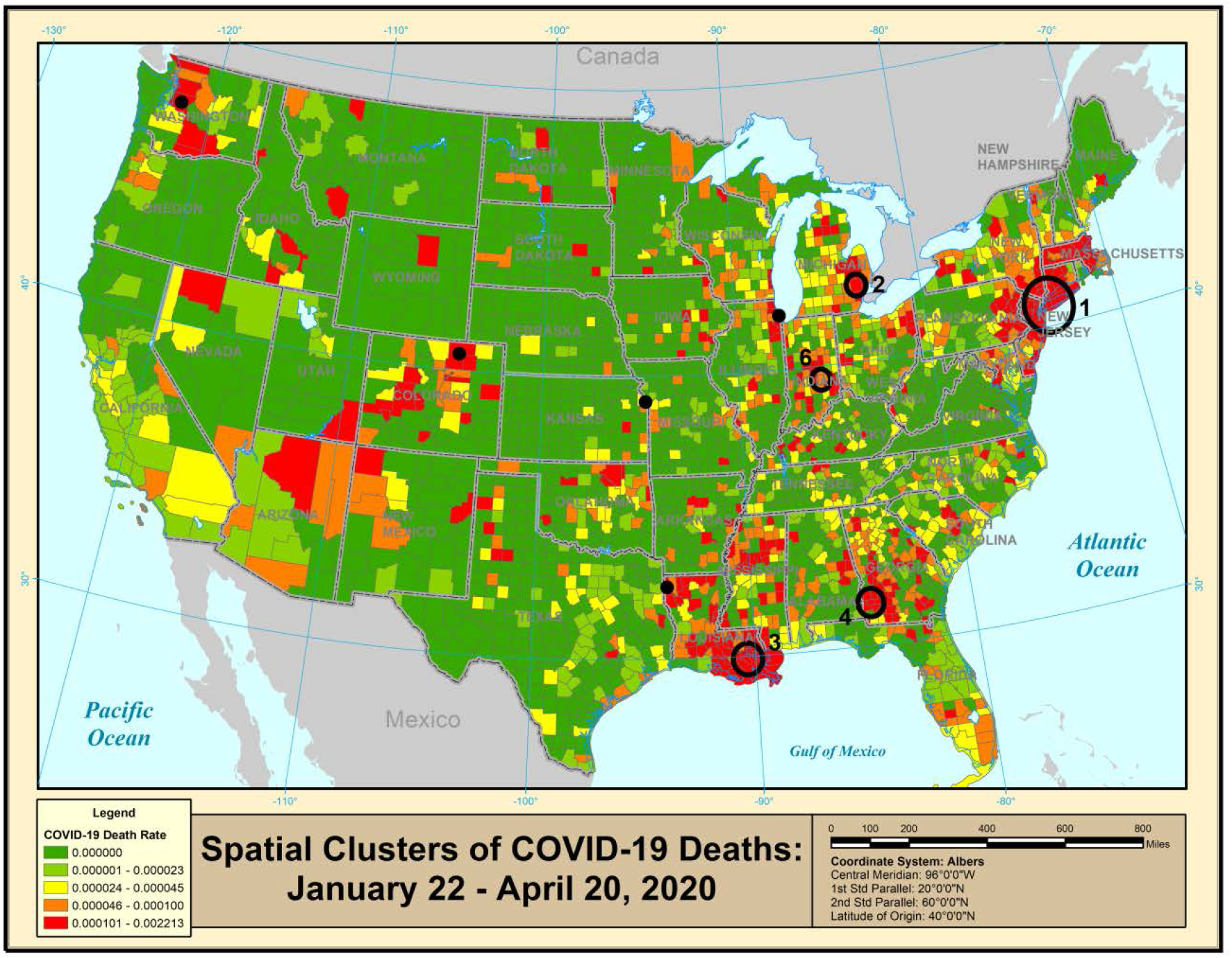
Geographical distribution and clusters of COVID-19 deaths for January 22-April 20, 2020.

**Table 2:** Statistically significant spatial clusters (p<0.05) of COVID-19 deaths for January 22-April 20, 2020, adjusted for age and multiple testing.

| Cluster Number | Locations | Relative Risk | Population |
| --- | --- | --- | --- |
| 1 | New York, New Jersey, Connecticut | 14.2 | 20,979,275 |
| 2 | Michigan (Detroit) | 4.7 | 3,880,899 |
| 3 | Louisiana (New Orleans) | 6.3 | 1,163,888 |
| 4 | Georgia (Atlanta) | 8.9 | 185,388 |
| 5 | Cook County, IL (Chicago) | 1.6 | 5,150,233 |
| 6 | Indiana (Indianapolis) | 1.9 | 1,372,565 |
| 7 | King County, WA (Seattle) | 1.6 | 2,252,782 |
| 8 | Caddo Parish, LA (Shreveport) | 2.5 | 240,204 |
| 9 | Weld County, CO (Greeley) | 2.4 | 324,492 |
| 10 | Woodson County, KS (Yates Center) | 2.9 | 165,429 |

### Recent Situation

For public health action, the more recent situation is of great interest. For the 14 days period April 7-20, 2020, there were 43 statistically significant clusters with a high rate of diagnosed cases, as shown in Figure 3. Table 3 provides details about the top 20 statistically significant clusters. The New York City metropolitan area is still the top cluster, while Marion, Ohio; Chicago; and New Orleans are number 2, 3 and 4 respectively. For deaths during the same period, there are nine significant clusters, as shown in Figure 4 and Table 4. The New York cluster is ranked number 1, with RR=13.5.

**Figure 3:**
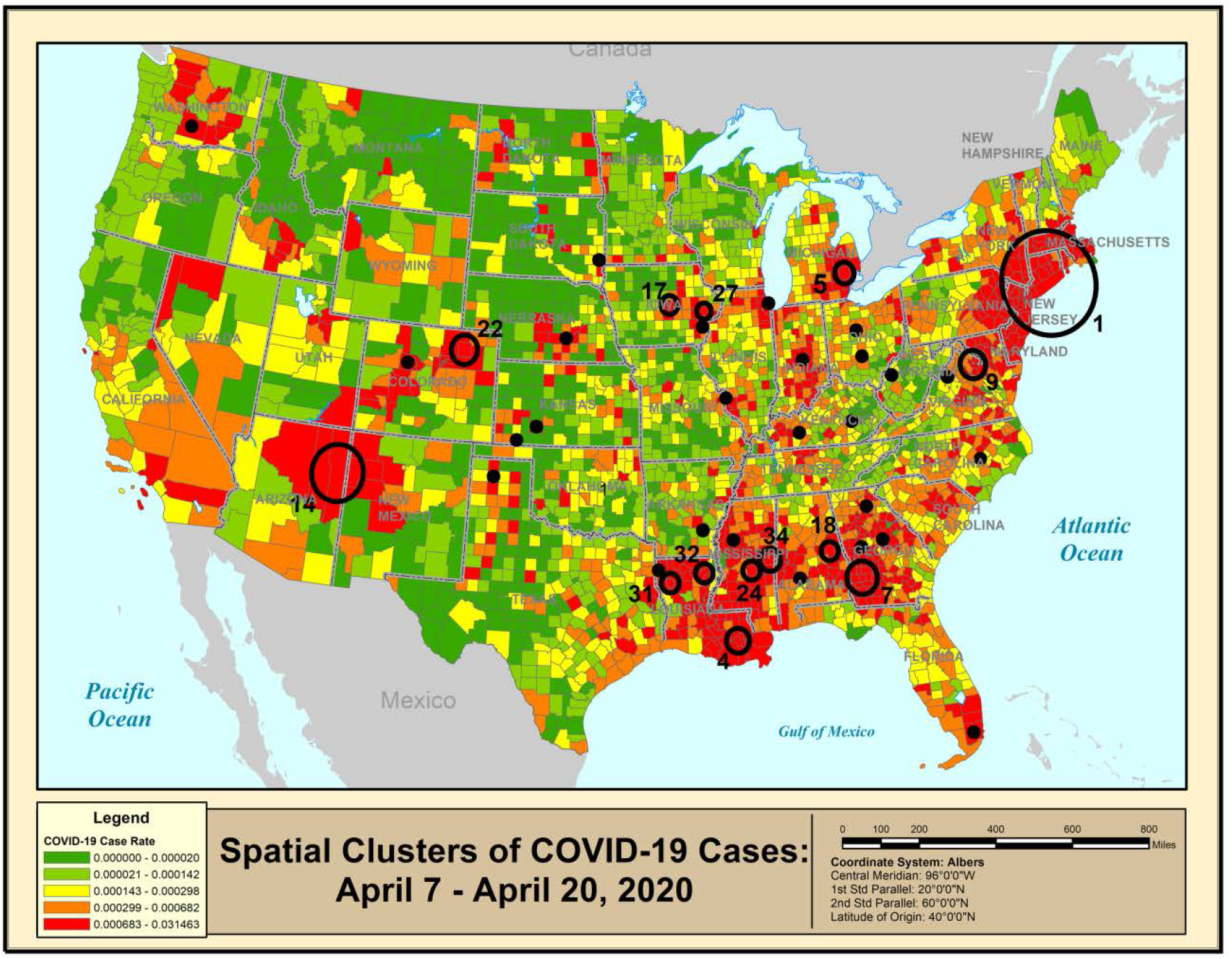
Geographical distribution and clusters of diagnosed COVID-19 cases for April 7-April 20, 2020.

**Figure 4:**
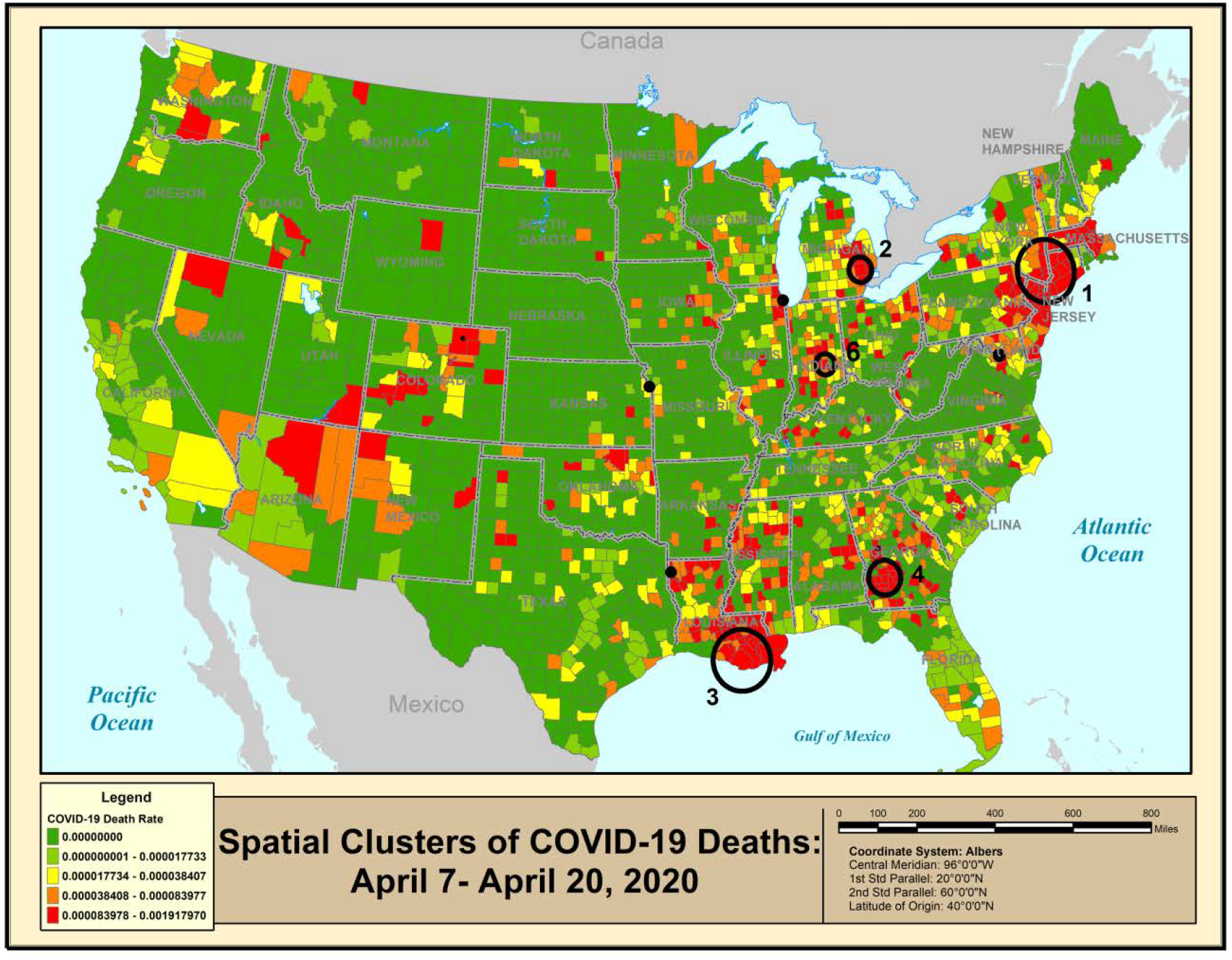
Geographical distribution and clusters of COVID-19 deaths for April 7-April 20, 2020.

**Table 3:** Statistically significant Spatial Clusters (p<0.05) of Age Adjusted COVID-19 Cases for April 7-April 20, 2020, adjusted for age and multiple testing.

| Cluster Number | Locations | Relative Risk | Population |
| --- | --- | --- | --- |
| 1 | New York, New Jersey, Connecticut, Pennsylvania, Rhode Island, Massachusetts | 8.4 | 31,835,482 |
| 2 | Marion County, OH (Marion) | 24.0 | 65,093 |
| 3 | Cook County, IL (Chicago) | 2.1 | 5,150,233 |
| 4 | Louisiana (New Orleans) | 3.6 | 1,037,284 |
| 5 | Michigan (Detroit) | 2.2 | 3,880,899 |
| 6 | Pickaway County, OH (Circleville) | 16.3 | 58,457 |
| 7 | Georgia (Atlanta) | 5.3 | 274,657 |
| 8 | Minnehaha County, SD (Sioux Falls) | 5.3 | 193,134 |
| 9 | Virginia, Maryland, Washington D.C. | 1.5 | 4,500,170 |
| 10 | Lincoln County, AR (Star City) | 14.5 | 13,024 |
| 11 | Louisa County, IA (Louisa) | 14.4 | 11,035 |
| 12 | Marion County, Indiana (Indianapolis) | 1.8 | 964,582 |
| 13 | Wayne County, NC (Goldsboro) | 3.6 | 123,131 |
| 14 | Arizona, New Mexico | 2.7 | 254,178 |
| 15 | Miami-Dade County, FL | 1.4 | 2,716,940 |
| 16 | Hall County, Nebraska (Grand Island) | 4.6 | 61,353 |
| 17 | Tama and Marshall Counties, IA (Marshall) | 4.5 | 56,223 |
| 18 | Chambers and Tallapoosa Counties, AL<br>(Alexander City) | 3.2 | 73,621 |
| 19 | Hall County, GA (Gainesville) | 2.3 | 204,441 |
| 20 | Harrisonburg City, VA | 3.9 | 53,016 |

**Table 4:** Statistically significant spatial clusters (p<0.05) of COVID-19 deaths for April 7-April 20, 2020, adjusted for age and multiple testing.

| Cluster Number | Locations | Relative Risk | Population |
| --- | --- | --- | --- |
| 1 | New York, New Jersey, Connecticut | 13.5 | 22,633,803 |
| 2 | Michigan (Detroit) | 4.2 | 4,533,447 |
| 3 | Louisiana (New Orleans) | 4.3 | 1,454,727 |
| 4 | Georgia (Albany) | 5.5 | 287,311 |
| 5 | Cook County, IL (Chicago) | 1.8 | 5,150,233 |
| 6 | Indiana (Indianapolis) | 2.1 | 1,372,565 |
| 7 | Woodson County, KS (Yates Center) | 3.5 | 165,429 |
| 8 | Henrico County, VA (Greater Richmond Region) | 2.4 | 330,818 |
| 9 | Caddo Parish, LA (Shreveport) | 2.5 | 240,204 |

### Emerging Problem Areas

The purely spatial analysis does not provide information about temporal changes. When using the prospective space-time scan statistic, while adjusting for purely geographical variation, we found six statistically significant clusters of rapidly increasing rates of diagnosed COVID-19 cases, as shown in Figure 5 and Table 5. The top cluster is in the Upper Midwest, stretching from Kansas and Missouri in the south to the Canadian border. The second cluster consists of several states in the Mid-Atlantic regions. There are also four geographically smaller clusters of increased COVID-19 activity in and around New York City. Figure 6 and Table 6 shows the equivalent results for mortality. The top cluster with a rapid increase in mortality is in Southern New England, followed by a huge cluster consisting of most of the Midwest.

**Figure 5:**
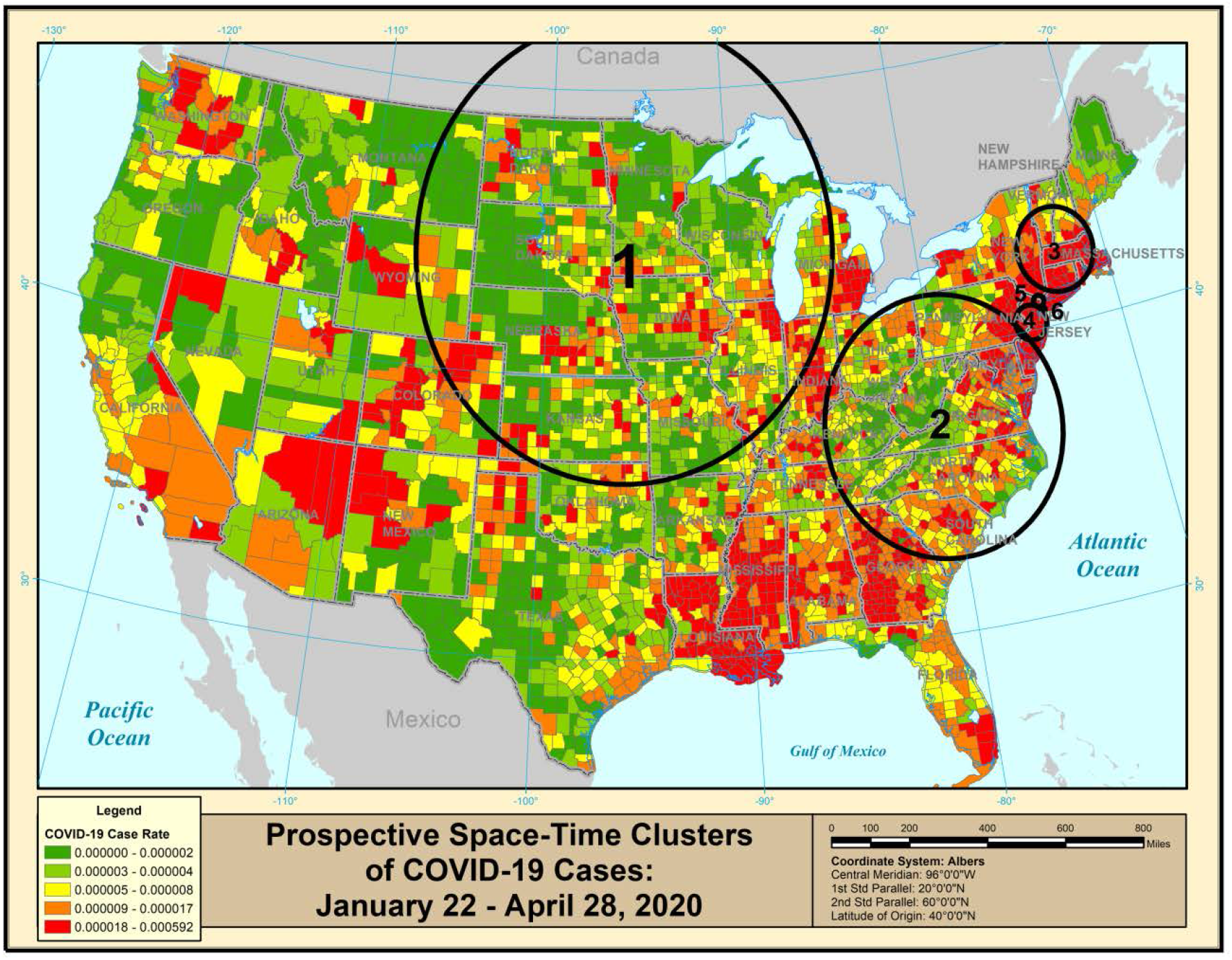
Geographical distribution and space-time clusters of diagnosed COVID-19 cases for January 22-April 28, 2020, with clusters adjusted for the geographical variation.

**Table 5:** Statistically significant space-time clusters of diagnosed COVID-19 cases for January 22-April 28, 2020, adjusted for geographical variation and multiple testing.

| Space-time Clusters of COVID-19 Cases: January 22-April 20, 2020 |  |  |  |
| --- | --- | --- | --- |
| Cluster Number | Locations | Relative Risk in Cluster | Cluster Population |
| 1 | Upper Midwest (MT, WY, CO, KA, OK, MO, IL, IN, MI, IA, NE, SD, ND, MN, WI) | 3.9 | 48,702,395 |
| 2 | Appalachian Region (PA, OH, KY, WV, MD, DE, VA, TN, NC, SC, GA, IN) | 3.8 | 59,455,758 |
| 3 | New England (MA, VT, NH, CT, RI, NY) | 3.7 | 14,559,398 |
| 4 | NJ (Mercer County, Middlesex County, Somerset County, Hunterdon County, Burlington County, Monmouth County, Union County, Camden County, Morris County, Ocean County, Essex County, Warren County) NY (Richmond County) PA (Bucks County, Philadelphia County, Montgomery County) | 2.9 | 9,295,418 |
| 5 | NJ (Bergen County, Passaic County) NY (Rockland County, New York County, Bronx County) | 2.4 | 4,806,730 |
| 6 | NY (Kings County, Queens County) | 2.2 | 4,813,761 |

**Figure 6:**
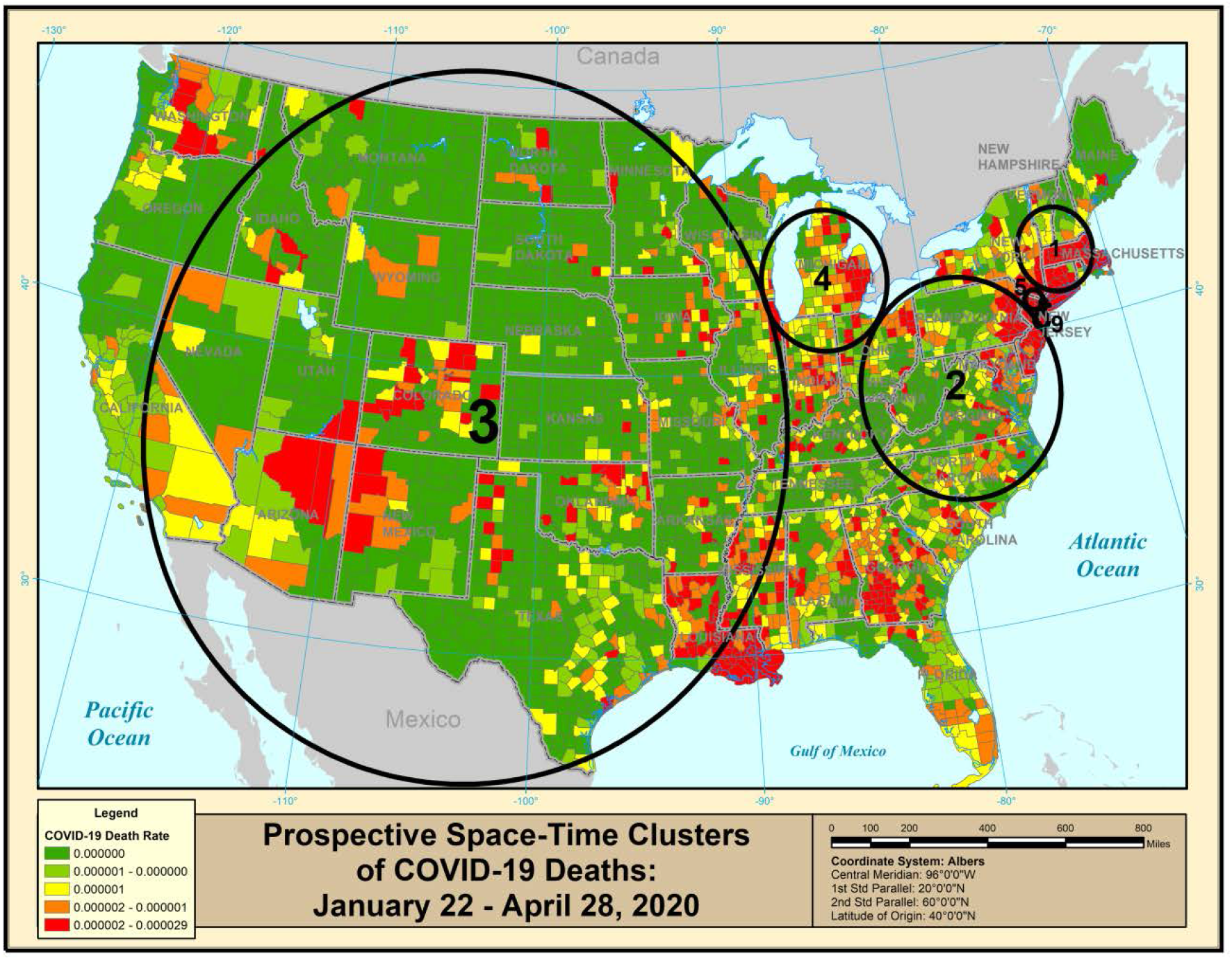
Geographical distribution and space-time clusters of COVID-19 deaths for January 22-April 28, 2020, with clusters adjusted for the geographical variation.

**Table 6:**
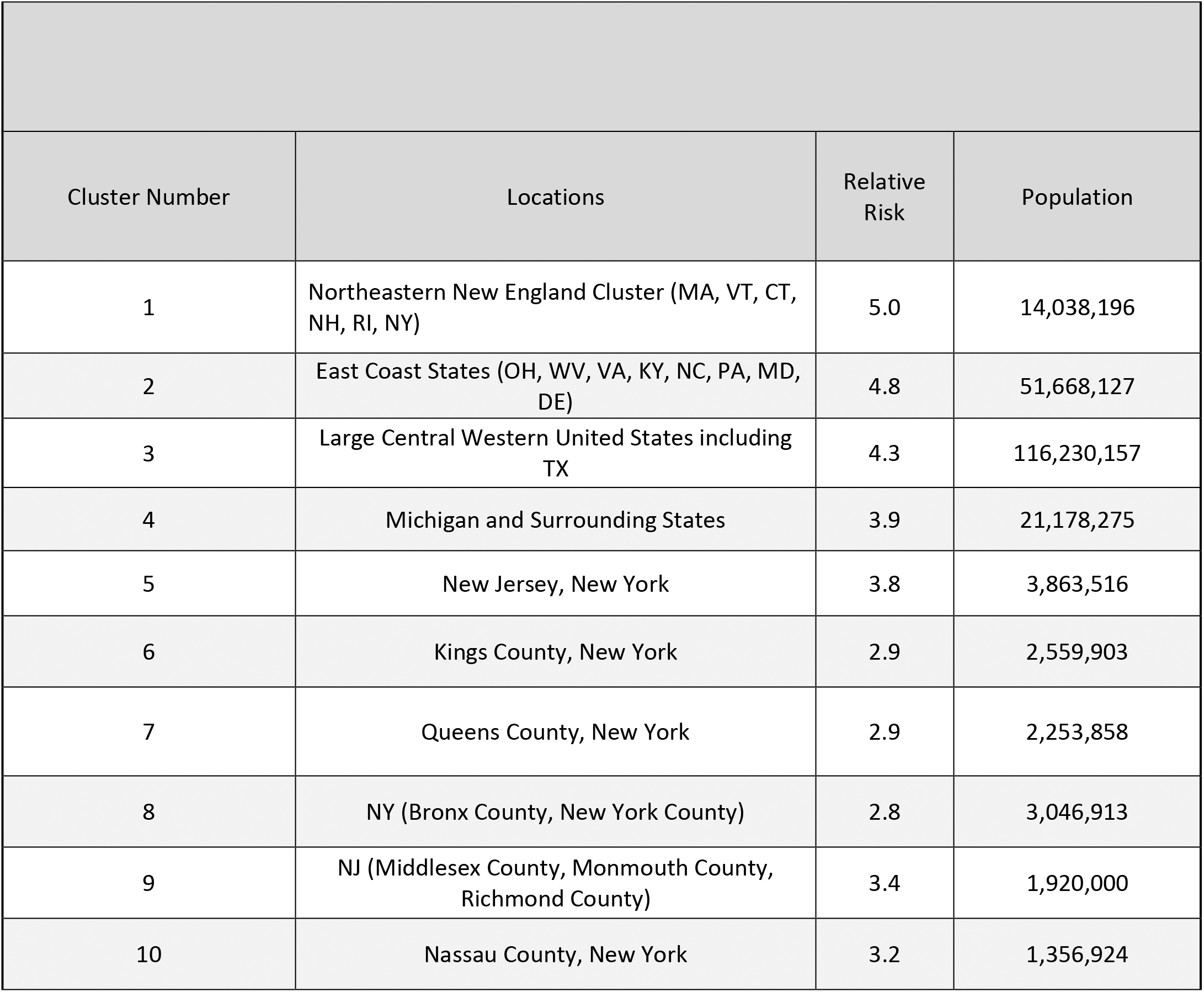
Statistically significant space-time clusters of COVID-19 deaths for January 22-April 28, 2020, adjusted for geographical variation and multiple testing.

## Discussion

To minimize COVID-19 mortality, it is important to monitor the spread of the disease in both space and time. Since older people have a much higher exposure and infection fatality ratio, it is important that they are protected and isolated until herd immunity is reached. For that, it is important both to know what places are closer to herd immunity and the places that currently have high infection and mortality rates.

The diagnosis and death data have different strength and weaknesses. The diagnosis data has the major weakness, in that the rates both depend on the presence of the disease as well as on the amount of testing. For a cluster to be detected, both are required. Hence, the analysis may have missed high incidence areas with low levels of testing. The mortality data is less dependent on testing, and it is reassuring that the two data sets produce somewhat similar results. One advantage of the diagnosis analyses is that it provides earlier indications about COVID-19 activity since the disease is typically diagnosed before death. An even earlier indication of COVID-19 activity may be syndromic data on influenza like illness, and spatial scan statistics have also been applied to such data. [12]

It is popular to compare COVID-19 mortality between countries and regions, but most such comparisons do not adjust for age, which is critical since COVID-19 is highly age dependent. It is also critical to note that some countries and regions are further along to reach herd immunity, and it is not necessarily the areas with fewer deaths that will have the fewest deaths by the time the pandemic is over.

This study applies modern disease surveillance methodology in addition to using choropleth maps that clearly show the relative ranking of counties’ virus cases and deaths. While the descriptive choropleth maps give an overall view of the contiguous US regarding the relative standings of counties for the COVID-19 virus levels, SaTScan™ provides the methodology to identify spatial clusters across the entire study period and also for the last 14 days, testing for significance of each identified cluster. Such spatial clusters show the geographic distribution of this virus when averaged across the study period. In addition to the spatial clusters, SaTScan™ also identified emerging clusters by using the prospective space-time analysis. These are outbreaks of COVID-19 cases and death within a window of 14 days and such information is helpful in understanding where in the contiguous US the virus is spreading the most.

While this study looked at COVID-19 across a large country, these types of analyses are equally or even more important to do for smaller regions in order to detect neighborhood clusters where the elderly require extra protection. That will require data at a finer geographical resolution available to many health departments but often publicly unavailable.

As COVID-19 is a rapid evolving pandemic, it is critical to have up to date information on its spread, of the type that cannot be provided by academic journals. As a complement to this paper, we will provide updated maps online. In near time, this will provide rapid up to date information regarding the United States. For the longer term, it will provide a historical record for how the disease spread over time and place.

## Conclusions

It is possible to quickly identify significant spatial clusters of COVID-19 cases and death based on county level data. However, different public health questions require different types of spatial statistical analyses. With properly focused analyses, this methodology can be useful to public health agencies to appropriately focus their COVID-19 counter measures.

## Data Availability

All COVID-19 data were downloaded from www.usafacts.org. All used data have been uploaded to https://github.com/jacobchurch/COVID-19_Geographical_Surveillance. All updates for this manuscript will be posted at this Git-Hub site.

## Declarations

### International novelty

The paper presents and illustrates different geographical analysis methods for monitoring COVID-19, explaining their different purposes and interpretation. The methods can be used in any country, and for national, regional and/or local data of different geographical resolution.

### Ethics approval and consent to participate

Not applicable

### Consent for publication

Not applicable

### Availability of data and materials

All COVID-19 data were downloaded from www.usafacts.org. All used data have been uploaded to https://github.com/iacobchurch/COVID-19_Geographical_Surveillance. All updates for this manuscript will be posted at this Git-Hub site.

## Authors’ contributions

RA and MK conceptualized the project and directed the modeling and the statistical data analyses. TH, JC and DS downloaded and analyzed the data with SaTScan and ArcMap, and generated the maps and tables. RA and MK wrote the manuscript. All authors edited and approved the final manuscript.

## References

1. Kamel Boulos MN and Geraghty EM. Geographical tracking and mapping of coronavirus disease COVID-19/severe acute respiratory syndrome coronavirus 2 (SARS-CoV-2) epidemic and associated events around the world: how 21st century GIS technologies are supporting the global fight against outbreaks and epidemics. Int J Health Geogr. 2020 Mar 11;19(1):8. doi: 10.1186/s12942-020-00202-8.

2. Johns Hopkins. CSSE Coronavirus COVID-19 Global Caaes (dashboard). https://gisanddata.maps.arcgis.com/apps/opsdashboard/index.html#/bda7594740fd40299423467b48e9ecf6.

3. World Health Organization. Novel coronavirus (COVID-19) situation (public dashboard). https://gisanddata.maps.arcgis.com/apps/opsdashboard/index.html#/bda7594740fd40299423467b48e9ecf6

4. Desjardins MR, Hohl A, Delmelle EM. Rapid surveillance of COVID-19 in the United States using a prospective space-time scan statistic: Detecting and evaluating emerging clusters. Applied Geography (Sevenoaks, England). 2020;118:102202

5. Kulldorff M. and Information Management Services, Inc. SaTScanTM v8.0: Software for the spatial and space-time scan statistics. http://www.satscan.org/, 2009.

6. https://usafacts.org/issues/coronavirus/

7. Centers for Disease Control and Prevention (cdc.gov) 1600 Clifton Rd., Atlanta, GA 30333.

8. U.S. Census Bureau. https://www.census.gov/data/datasets/time-series/demo/popest/2010s-counties-total.html#par_textimage_70769902

9. Centers for Disease Control and Prevention. Severe Outcomes Among Patients with Coronavirus Disease 2019 (COVID-19) — United States, February 12–March 16, 2020. CDC COVID-19 Response Team. US Department of Health and Human Services/Centers for Disease Control and Prevention MMWR / March 27, 2020 / Vol. 69 / No. 12

10. Kulldorff M. A spatial scan statistic. Communications in Statistics: Theory and Methods, 1997; 26: 1481–1496.

11. Kulldorff M. Prospective time periodic geographical disease surveillance using a scan statistic. Journal of the Royal Statistical Society: Series A (Statistics in Society). 2001;164(1):61–72.

12. Aledade Inc, Kulldorff M. COVID-19 syndromic survey geographic clusters (data through 05–12-2020), https://covidmap.aledade.com/survey-symptom-clusters/, 2020.

